# Plasma soluble TREM2 is associated with plasma pTau-181 and pTau-231 in cognitively normal older adults at risk of Alzheimer’s disease

**DOI:** 10.1101/2025.08.11.25333037

**Authors:** Prita R. Asih, Cameron W. Morris, Hong Wang, Steve Pedrini, Kathryn Goozee, Simon M. Laws, Pratishtha Chatterjee, Kevin Taddei, Hamid R. Sohrabi, Stephanie R Rainey-Smith, Chai K. Lim, Gilles J. Guillemin, Thomas K. Karikari, Colin L. Masters, Henrik Zetterberg, Kaj Blennow, Nicholas J. Ashton, Ralph N. Martins

**Affiliations:** Macquarie Medical School, Faculty of Medicine, Health and Human Sciences, Macquarie University, Sydney, NSW 2109, Australia; Alzheimer Research Australia, Sarich Neuroscience Research Institute, Nedlands, WA, Australia; Lilly Research Laboratories, Eli Lilly and Company, Indianapolis, IN, USA; School of Medical Health and Sciences, Edith Cowan University, Joondalup, WA, Australia; KaRa Institute of Neurological Disease, Sydney, Macquarie Park, Australia; Centre for Precision Health, Edith Cowan University, Joondalup, WA, Australia; Collaborative Genomics and Translation Group, School of Medical and Health Sciences, Edith Cowan University, Joondalup, Western Australia, Australia; Curtin Medical School, Curtin University, Bentley, Australia; Florey Institute of Neuroscience and Mental Health, University of Melbourne, Parkville, Victoria, Australia; Centre for Healthy Ageing, Health Future Institute, Murdoch University, Murdoch, Western Australia, Australia; Department of Psychiatry and Neurochemistry, Institute of Neuroscience and Physiology, the Sahlgrenska Academy at the University of Gothenburg, Mölndal, Sweden; Clinical Neurochemistry Laboratory, Sahlgrenska University Hospital, Mölndal, Sweden; Department of Neurodegenerative Disease, UCL Institute of Neurology, London, UK; UK Dementia Research Institute at UCL, London, UK; Hong Kong Center for Neurodegenerative Diseases, Hong Kong, China; Wisconsin Alzheimer’s Disease Research Center, University of Wisconsin School of Medicine and Public Health, University of Wisconsin-Madison, Madison, Wisconsin, USA; Institute of Neuroscience and Physiology, University of Gothenburg, Mölndal, Sweden; Paris Brain Institute, ICM, Pitié-Salpêtrière Hospital, Sorbonne University, Paris, France; Neurodegenerative Disorder Research Center, Division of Life Sciences and Medicine, and Department of Neurology, Institute on Aging and Brain Disorders, University of Science and Technology of China and First Affiliated Hospital of USTC, Hefei, Anhui, P.R. China; King’s College London, Institute of Psychiatry, Psychology and Neuroscience, Maurice Wohl Institute Clinical Neuroscience Institute, London, UK; NIHR Biomedical Research Centre for Mental Health and Biomedical Research Unit for Dementia at South London and Maudsley NHS Foundation, London, UK; Centre for Age-Related Medicine, Stavanger University Hospital, Stavanger, Norway; Senior Director, Banner Research Blood-Based Biomarker Program, Banner Health, Phoenix, AZ, United States; Department of Neuroscience, Monash University, Melbourne, Victoria, Australia; Bionyeri Pty Ltd, Hornsby, NSW Australia; Department of Psychiatry, University of Pittsburgh, Pittsburgh, PA 15213, USA; Department of Neurology, Duke University Medical Center, Durham, North Carolina, USA; Department of Chemistry.Faculty of Mathematics and Natural Sciences. IPB University, Bogor, Indonesia

**Keywords:** plasma sTREM2, plasma p-tau, biomarkers, microglia activation, neuroinflammation, Alzheimer’s disease, R47H TREM2

## Abstract

**Background:** CSF and blood soluble TREM2 (sTREM2) levels have been found to increase at early stage of Alzheimer’s Disease (AD). The relationships between sTREM2, AD-related biomarkers, and other neuroinflammation biomarkers remain unclear. Moreover, the impact of rare variants in TREM2 gene (R47H/R62H), which are associated with increased risk of AD, on plasma sTREM2 has not been elucidated.

**Objective:** Investigate the association of plasma sTREM2 levels with brain Aβ load and AD-related blood biomarkers such as phosphorylated tau (pTau)-181, pTau-231, GFAP, NFL, and other neuroinflammation and peripheral inflammation markers in cognitively normal (CN) older adults at risk of AD (CN Aβ+) compared to CN Aβ-, including the effect of AD-linked TREM2 rare variants.

**Methods:** Plasma sTREM2 concentrations were measured by MesoScale Discovery (MSD) assay from the KARVIAH cohort. Participants underwent cognitive tests and PET amyloid imaging. Genetic data and blood biomarkers were included for correlation analysis. Associations with plasma sTREM2 were investigated upon stratification by PET-Aβ load SUVR ((CN Aβ- (n=65) and CN Aβ+ (n=35)) as the main analysis. A subgroup analysis based on the *TREM2 R47H* genotype was carried out as exploratory analysis.

**Results:** Plasma sTREM2 positively correlated with plasma pTau181, and pTau231 in CN Aβ+ group. Plasma sTREM2 positively correlated with serum microglial kynurenine pathway metabolites. Plasma sTREM2 and brain Aβ load were higher in R47H TREM2 carriers compared to non-carriers.

**Conclusions:** Plasma sTREM2, a marker of microglia activation, is associated with plasma pTau181 and 231 in cognitively normal older adults at risk of AD, with higher plasma sTREM2 levels in R47H TREM2 carriers.

## INTRODUCTION

Triggering receptor expressed on myeloid cells 2 (TREM2) is a type 1 transmembrane protein expressed in cells of the myeloid lineage (the progenitors of microglia), and its rare variant of *TREM2* gene – the R47H variant - has been shown to substantially increase the risk of developing Alzheimer’s disease (AD) by ∼3-4 fold ^1^. *TREM2* R47H poses a risk for AD that is similar to that conferred by one copy of the Apolipoprotein (*APOE)* ε4 allele ^2^. This strong association with AD supports the active role of immune and inflammatory pathways as a potential cause of the disease, rather than as a consequence ^3^. Evidence from transgenic mouse models showed that R47H variant and other *TREM2* AD-associated variants such as R62H caused partial loss of function of the TREM2 protein and modify the nature of microglia in response to amyloid-β (Aβ) plaques ^4, 5^. Because of the low frequency (0.2%) of the R47H variant of *TREM2*, people homozygous for this variant are extremely rare ^6^.

TREM2 is highly and exclusively expressed by microglia in the brain ^2^ and myeloid cells in the blood, and its ectodomain binds to apolipoproteins, phospholipids, lipoproteins, Aβ, and galectin-3 ^7–9^. The surface levels of TREM2 are regulated via cleavage by ADAM10 and ADAM17 that generate soluble fragment (sTREM2) which is released into the extracellular space and can be detected in CSF and plasma ^10, 11^. The intramembrane and transmembrane domains of the remaining stalk are subsequently cleaved by γ-secretase activity ^12^. Around 25% of sTREM2 production originates from the alternative splicing of TREM2 lacking the transmembrane domain ^12^.

The potential application of sTREM2 as a putative neuroinflammatory marker for AD has been extensively investigated, particularly as a marker for microglia activity in the early stage of the disease ^13–15^. CSF sTREM2 levels have been found to be increased with age ^16^, and to be elevated in autosomal dominant AD patients ^17, 18^ and in early stages of sporadic AD ^10, 13, 19, 20^. Moreover, CSF sTREM2 levels have been shown to be affected by the *TREM2* genetic variants ^10^, for example R47H carriers had higher CSF sTREM2 ^20^. Interestingly, Aβ and tau pathology may affect CSF sTREM2 regulation differently, such that sTREM2 levels have been shown to correlate with those of CSF total tau and phospho-tau but not with Aβ42 levels ^16, 19, 20^. Furthermore, increased CSF sTREM2 levels in autosomal dominant AD patients may occur approximately 5 years before symptoms onset and after increased levels of Aβ deposition and CSF tau levels ^17^.

While studies highlight CSF sTREM2 levels as a biomarker for AD diagnosis or progression, the invasive nature of CSF collection motivated the investigation of sTREM2 in a more accessible blood-based biofluids (serum and plasma). To date, six studies have assessed sTREM2 levels in blood; however, due to conflicting results, the utility of sTREM2 as a peripheral biomarker for AD remains unclear. Specifically, some have found no difference in sTREM2 plasma/serum levels between healthy controls, AD patients ^10, 11, 21, 22^, or *TREM2* variant carriers ^11^, suggesting only CSF sTREM2 levels, not plasma, is an informative AD biomarker. In contrast, three studies highlight the utility of blood/serum sTREM2 as an AD biomarker, as evidenced by increased levels in blood from AD patients ^23^ and a positive association with the risk of dementia ^15, 22^. While these studies hypothesize blood sTREM2 as a candidate biomarker for AD, it remains unclear if plasma sTREM2 levels can serve as a biomarker for AD progression, such as in pre-clinical AD (characterized by Neocortical Aβ Load or brain Aβ load).

To help address these inconsistencies, this study examines plasma sTREM2 levels in cognitively normal older adults at risk for AD. Given that aberrant neocortical Aβ load accumulation starts as early as two decades before the clinical manifestation ^24^, we first investigated the association between plasma sTREM2 levels and brain Aβ load as the main analysis. Furthermore, to better understand how plasma sTREM2 levels change for known AD pathophysiology, we also investigated the relationship between plasma sTREM2 with standard AD blood biomarkers such as phosphorylated tau (pTau), neurofilament light, (NFL), glial fibrillary acidic protein (GFAP), and additional neuroinflammation markers of kynurenine pathway (KP) metabolites and peripheral inflammation markers of C-reactive protein as the main analysis. Lastly, we further assessed how this association is impacted by the AD genetic risk factors, the *TREM2* R47H/R62H rare variants as an exploratory analysis.

## METHODS

### Participants

Study participants belonged to the Kerr Anglican Retirement Village Initiative in Ageing Health (KARVIAH) cohort, at baseline. All participants were residents of Anglicare, New South Wales, Australia. Cohort volunteers (N = 206) were required to meet the set screening inclusion and exclusion criteria to be eligible for the KARVIAH cohort. Briefly, the inclusion criteria for the KARVIAH cohort comprised an age range of 65–90 years, good general health, no known significant cerebral vascular disease, fluent in English, adequate/corrected vision and hearing to enable testing, no objective cognitive impairment as screened by the Montreal Cognitive Assessment (MoCA) cut off score ≥ 26. The MoCA scores lying between 18–25 were assessed on a case by case basis by the study neuropsychologist following stratification of scores according to age and education ^25^. The exclusion criteria comprised, the diagnosis of dementia based on the revised criteria from the National Institute on Aging - Alzheimer’s Association ^26^, presence of acute functional psychiatric disorder (including lifetime history of schizophrenia or bipolar disorder), history of stroke, untreated severe or extremely severe depression (based on the depression, anxiety, stress scales; DASS) and uncontrolled hypertension (systolic BP > 170mm Hg or diastolic BP > 100mm Hg).

One hundred and five participants out of the 134 volunteers meeting the inclusion/exclusion criteria, underwent neuroimaging, neuropsychometric evaluation and blood collection, as the remaining either declined undergoing neuroimaging or withdrew from the study. Within these 105 participants, 100 participants were considered to have un-impaired global cognition based on their Mini-Mental State Examination score ^27^ (MMSE≥26) and were included in the current study. Plasma sTREM2 concentrations were reported in all 100 participants considered to have normal global cognition. All volunteers provided written informed consent prior to participation, and the Bellberry Human Research Ethics Committee, Australia, provided approval for the study. Details of the participants included within the current study have been illustrated in **Supplementary Figure 1**.

### Neuroimaging assessments and brain Aβ classification

All study participants were imaged within three months of blood collection. Participants underwent positron emission tomography (PET) imaging using ligand ^18^F-Florbetaben (FBB) at Macquarie Medical Imaging in Sydney. Participants were administered an intravenous bolus of FBB slowly over 30 s, while in a rested position. Images were acquired over a 20 min scan, in 5 min acquisitions, beginning 50 min post injection. The mean standard uptake value ratio, SUVR, of various neocortical regions, containing the (i) frontal, (ii) superior parietal, (iii) lateral temporal, (iv) lateral occipital, and (v) anterior and posterior cingulate, were measured employing CapAIBL, an image processing software, to estimate the brain Aβ load^28, 29^. We employed a cut-off value of 1.35, categorizing study participants as CN Aβ+ (SUVR≥1.35) and CN Aβ- (SUVR<1.35), as published previously ^30–32^. The cerebellar cortex was used as the reference region for SUVR quantification.

### Blood collection, APOE, TREM2 R47H and R62H genotyping, measurement of plasma sTREM2 and other AD blood biomarkers

All study participants fasted for a minimum of 10 hours overnight prior to blood withdraw employing standard serological methods and processing ^30^. The *APOE* genotype was determined from purified genomic DNA extracted from 0.5 ml whole blood as previously described ^30^. Then, each sample was genotyped for the presence of the R47H and R62H (the two most common AD-associated variants of *TREM2*) based on Taqman SNP genotyping assays for rs75932628 (Assay ID C_100657057_10) and rs143332484 (Assay ID C_172216876_10), respectively, as per the manufacturer’s instructions (AB Applied Biosystems by Life Technologies, Scoresby, VIC, Australia). Since the rare missense R47H variant in *TREM2* has been shown to mediate LOAD risk substantially in Icelandic and Caucasian populations ^33^, we chose to investigate the effect of this variant in our KARVIAH cohort which comprises mostly Anglo-Saxon or Caucasian. Ninety percent of the samples were genotyped in singlicate and five percent in duplicate, and 100% inter- and intra-assay concordance was observed. Plasma sTREM2 concentrations were measured employing the Meso Scale Discovery (MSD) assay developed at Eli Lilly and Company using rabbit monoclonal antibodies generated against the extracellular domain of human TREM2. HSA-tagged human TREM2 (hTREM2 19-174) recombinant protein was used as standard. All plasma samples were run at a 1:64 dilution. Analyses of KP metabolites were performed simultaneously with ultra-high performance liquid chromatography (UHPLC) with an injection volume of 20 μL per sample, as described previously ^31^. Plasma NFL was measured using the ultra-sensitive single-molecule array (Simoa) platform as previously described ^32^. Plasma GFAP and total Tau (tTau) were measured using the Neurology 4-Plex A kit (QTX-102153, Quanterix, Billerica, United States) at Edith Cowan University, Perth, Australia whereas plasma pTau181 and pTau231 were measured using the in-house assay developed at the University of Gothenburg, Sweden, as previously described ^34^.

### Statistical analyses

Cohort characteristics were reported as means + SD (continuous variables) and percentage values (categorical variables). Independent sample *t*-tests or Chi-square tests were employed to compare the sex, the frequency of *APOE* ε4, status, and plasma sTREM2 levels between CN Aβ+ and CN Aβ-groups. Fisher’s exact test was employed to determine TREM2 R47H and R62H carrier frequency differences. Continuous response variables were tested for approximate normality and variance homogeneity using the Kolmogorov-Smirnov test and the Shapiro-Wilk test, and log-transformed when required to approximate a better normal distribution. After transformation, the data followed a normal distribution. All data analysis reported has been performed on log_10_-transformed sTREM2, but the untransformed values are shown in descriptive tables and figures. Spearman correlation coefficient (*p*) was employed to investigate correlations between sTREM2 and other continuous variables of interest in all participants, CN Aβ-, and CN Aβ+ group. Additionally, linear models were utilised to calculate correlations after adjusting for confounding variables, including age, sex, and *APOE* ε 4 status, followed by false discovery rate (FDR) adjustment for correcting multiple comparisons; *p*-values < 0.05 that survived FDR adjustment were considered significant. All analyses were carried out using IBM® SPSS® Version 28. Figures were built using GraphPad Prism version 9.2.0. All tests were 2-tailed, with a significance level of α = 0.05.

## RESULTS

### Cohort characteristics

The cohort participant characteristics, including demographic, PET brain Aβ load, and AD-related blood biomarkers are presented in **Table 1**. As the demographic factors reported previously^31, 32^, the cohort differed significantly between the CN Aβ− and CN Aβ+ groups for the brain Aβ load (FBB-PET) and *APOE* ε4 carrier status as expected; however, no statistically significant differences were noted for age, sex, body mass index (BMI), and education status. The cohort had significant differences in plasma pTau181, pTau231, GFAP, and some of the kynurenine pathway (KP) metabolites such as kynurenine and anthranilic acid; however, no statistically significant differences in plasma total Tau, NFL, the remaining KP metabolites, and C-reactive protein (CRP). Interestingly, the frequency of the rare heterozygous R47H carrier was only found in the CN Aβ+ group. No significant differences were found between the study groups for cognitive measures and hippocampal volume^31,32^.

**Table 1.** Cohort characteristic. Baseline characteristics including demographic and plasma measures of study participants. *Note*: Categorical measures are presented as counts and percentages, whereas continuous measures are 11 presented as mean + SD. Independent sample *t*-tests, chi-square tests or Fisher’s exact tests were used 12 to compare the measures between the CN Aβ− and CN Aβ+ groups. All values are presented up to three 13 decimal places for the complete table, except for the <0.0001 values, and p-values< 0.05 were considered statistically significant (bold font). PET-Aβ load was categorised based on the FBB-1 PETSUVR cut-off of 1.35. *n* represents the count. All *p*-values for plasma pTau181, pTau231, 2 neurofilament light chain (NFL), and the kynurenine pathway (KP) metabolites were obtained from 3 variables transformed to the logarithmic scale for analyses to meet assumptions of the statistical test 4 employed. Abbreviations: APOE, apolipoprotein E; FBB-Aβ PET,18F-florbetaben positron emission 5 tomography; GFAP, Glial Fibrillary Acidic Protein; SUVR, standardised uptake value ratio.

| | All participants | CN Aβ<sup>-</sup> (n=65) | CN Aβ<sup>+</sup> (n=35) | Effect size (d) | p-value | 95% CI |
| --- | --- | --- | --- | --- | --- | --- |
| Sex (M/F) | 32/68 | 19/46 | 13/22 | 0.263 | 0.419 | -0.15-0.675 |
| Age (years) | 78.18 ± 5.52 | 77.61 ± 5.55 | 79.22 ± 5.38 | -0.294 | 0.165 | -0.706-0.12 |
| Body mass index (BMI) | 27.62 ± 4.48 | 27.42 ± 4.47 | 27.98 ± 4.55 | -0.231 | 0.553 | -0.564-0.157 |
| Education (years) | 14.43 ± 3.26 | 14.84 ± 3.37 | 13.64 ± 2.91 | -0.352 | 0.078 | -0.642-0.127 |
| n APOE ε4 carriers (%) | 21 (21) | 5 (7.69) | 16 (45.71) | -0.961 | **<0.0001** | -1.391-(-0.526) |
| n TREM2 R47H heterozygous carriers (%) | 5 (5) | 0 (0) | 5 (14.28) | 0.683 | **0.004** | 0.26-1.103 |
| n TREM2 R62H heterozygous carriers (%) | 3(3) | 2 (3.07) | 1 (2.86) | 0.244 | 0.28 | -0.169-0.656 |
| FBB-Aβ PET (SUVR) | 1.35 ± 0.31 | 1.15 ± 0.08 | 1.71 ± 0.26 | -3.578 | **<0.0001** | -4.221-(-2.926) |
| Total Tau; tTau (pg/ml) | 1.27 ± 0.38 | 1.2 ± 0.36 | 1.35 ± 0.4 | -0.311 | 0.065 | -0.728-0.108 |
| pTau181 (pg/ml) | 15.53 ± 5.64 | 13.55 ± 5.61 | 17.52 ± 5.68 | -0.726 | **0.002** | -1.161-(-0.287) |
| pTau231 (pg/ml) | 15.39 ± 6.92 | 11.57 ± 6.44 | 19.21 ± 7.39 | -1.057 | **2.00E-06** | -1.51-(-0.6) |
| GFAP (pg/ml) | 179.17 ± 68.01 | 146.96 ± 49.98 | 211.39 ± 86.04 | -1.048 | **7.00E-06** | -1.487-(-0.604) |
| Neurofilament Light Chain; NFL (pg/ml) | 36.38 ± 17.78 | 35.23 ± 17.27 | 38.5 ± 18.76 | -0.225 | 0.384 | -0.642-0.194 |
| C-reactive protein (CRP) (mg/l) | 2.12 ± 2.5 | 2.14 ± 2.26 | 2.07 ± 2.92 | 0.4 | 0.925 | -0.02-0.818 |
| *Kynurenine Pathway (KP) metabolites* |  |  |  |  |  |  |
| Tryptophan (μM) | 43.33 ± 7.72 | 42.87 ± 8.15 | 44.17 ± 6.87 | -0.197 | 0.35 | -0.613-0.221 |
| Kynurenine (μM) | 2.23 ± 0.58 | 2.12 ± 0.52 | 2.41 ± 0.63 | -0.519 | **<0.05** | -0.939-(-0.096) |
| Kynurenic acid (nM) | 52.06 ± 27.44 | 49.71 ± 26.25 | 56.41 ± 29.4 | -0.16 | 0.238 | -0.577-0.256 |
| 3-Hydroxykynurenine (nM) | 119.45 ± 37.47 | 117.74 ± 34.73 | 122.6 ± 42.42 | -0.232 | 0.554 | -0.649-0.185 |
| 3-Hydroxyanthranilic acid (nM) | 22.63 ± 9.14 | 22.15 ± 9.25 | 23.5 ± 8.99 | -0.194 | 0.406 | -0.611-0.223 |
| Anthranilic acid (nM) | 35.48 ± 19.79 | 31.4 ± 14.99 | 43.04 ± 25.02 | -0.684 | **<0.005** | -1.109-(-0.255) |
| Picolinic acid (nM) | 109.38 ± 49.82 | 112.06 ± 56.25 | 104.4 ± 35.07 | 0.081 | 0.572 | -0.335-0.497 |
| Quinolinic acid (nM) | 171.83 ± 74.48 | 169.76 ± 75.89 | 175.66 ± 72.71 | -0.117 | 0.689 | -0.532-0.298 |

*Note:* Categorical measures are presented as counts and percentages, whereas continuous measures are presented as mean ± SD. Independent sample *t*-tests, chi-square tests or Fisher's exact tests were used to compare the measures between the CN Aβ <sup>-</sup> and CN Aβ <sup>+</sup> groups. All values are presented up to three decimal places for the complete table, except for the <0.0001 values, and p-values< 0.05 were

### Associations of AD-related genetic and demographic factors with plasma sTREM2

Plasma sTREM2 was observed to correlate with age (r=0.365, *p*=0.0002) and R47H carriage (mean ± SD: non-carriers, 27,499 ± 11,034 pg/ml; carriers, 39,381 ± 7,498 pg/ml, *p*=0.019), while no significant association between plasma sTREM2 was observed with education (r=0.296 *p*=0.175), sex (mean ± SD males, 29,963 ± 2,929 pg/mL; females, 28,161 ± 1,197 pg/mL; *p*=0.501), BMI (r=-0.057 *p*=0.565) or *APOE* ε4 carriage (mean ± SD: non-carriers, 28,632 ± 1,466 pg/mL; carriers, 29,236 ± 2,390 pg/mL; *p*=0.844). Furthermore, plasma sTREM2 was not observed to be significantly elevated in *APOE* ε4 carriers compared to non-*APOE* ε4 carriers (mean ± SD; *APOE* ε4, 32,390.891 ± 20,778.251 pg/mL; non-*APOE* ε4, 29,085.951 ± 13,544.021 pg/mL) with (*p*=0.477).

### Association between plasma sTREM2 and Brain A**β** Load (*SUVR*) based on brain Aβ status

Plasma sTREM2 was not observed to be significantly elevated in CN Aβ+ participants (pre-clinical AD or at risk of AD) compared to CN Aβ-participants (mean ± SD; CN Aβ-, 28,432 ± 1,701 pg/mL; CN Aβ+, 29,342 ± 1,767 pg/mL) with (*p*=0.546) and without (*p*=0.729) adjusting for age, sex, and *APOE* ε4 carrier status. Furthermore, plasma sTREM2 levels were also not found to be correlated with brain Aβ load in all participants, as well as in CN Aβ-participants, before and after adjusting for age, sex, and *APOE* ε4 (**Table 2**). Positive correlation was observed in CN Aβ+ participants before adjusting for age, sex, and *APOE* ε4 carrier status; however, this positive correlation disappeared after adjusting for covariates (**Table 2**).

**Table 2.**
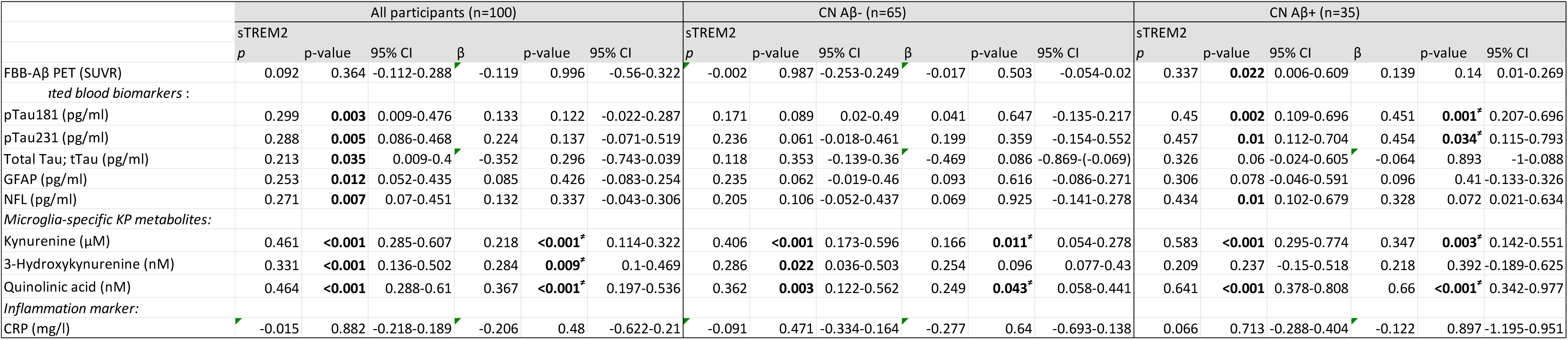
Plasma sTREM2 association with brain Aβ load, AD-related biomarkers, neuroinflammation, and peripheral inflammation markers in all cipants, CN Aβ- and CN Aβ+. *Note*: Plasma sTREM2 association with brain Aβ load, AD-related biomarkers, neuroinflammation, and peripheral inflammation markers were investigated using the Spearman 4 correlation, and *p*-values < 0.05 (bold) were considered significant. Generalised linear models were utilised to explore plasma sTREM2 association with brain Aβ load, AD-5 related biomarkers, neuroinflammation, and peripheral inflammation markers upon adjusting for age, gender, and *APOE* ε4 status, followed by FDR correction. *n*’ represents 6 the number of participants, ‘*ρ*’ represents the Spearman correlation coefficient and *β*’ represents the beta coefficient. Parameters significantly correlating (*p*-values < 0.05, bold) 7 with plasma sTREM2 after adjusting for confounding variables and survived FDR adjustment (*p*-values < 0.05, bold and ‘≠’ signed) were considered significant.

### Association between plasma sTREM2 and AD-related blood biomarkers based on brain Aβ status

AD-related blood biomarkers associated with plasma sTREM2 were investigated in all participants and separately in CN Aβ− and CN Aβ+ utilising the Spearman correlation coefficient and then generalised linear model accounting for age, sex, and *APOE* ε4 status followed by FDR correction (**Table 2**). In all participants, a significant positive correlation of plasma sTREM2 was observed with plasma total Tau, pTau181 and 231, GFAP, and NFL before adjusting for covariates, although after adjusting for covariates, these correlations did not remain significant (**Table 2**).

While investigating separately in CN Aβ− and CN Aβ+ individuals, a significant positive correlation of plasma sTREM2 was observed in CN Aβ+ participants with plasma pTau181, 231, and NFL before adjusting for covariates, and plasma sTREM2 remained significantly correlated with pTau181 and 231 after adjusting for covariates (**Table 2**, **Figure 1**).

**Figure 1.**
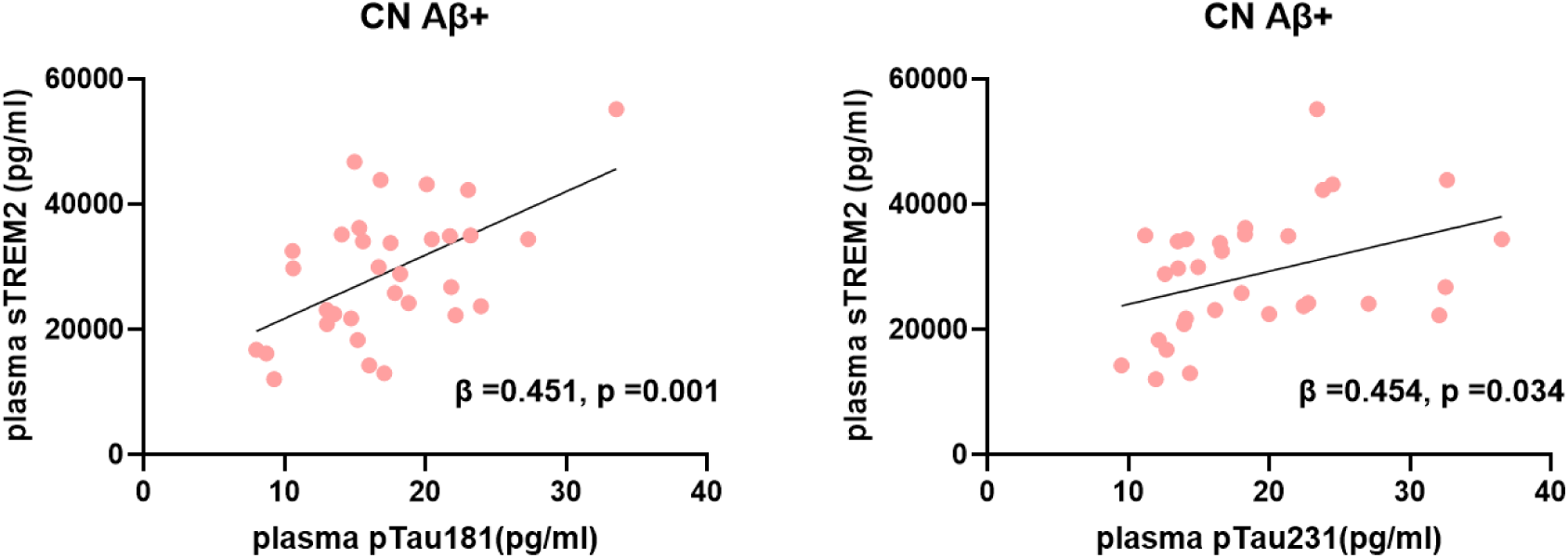
Association between plasma sTREM2 with plasma pTau (181 and 231). AD-related blood biomarkers significantly correlating with plasma sTREM2 both before and after adjusting for confounding variables and survived false discovery rate correction have been reported. Plasma sTREM2 concentrations (pg/ml) positively correlated with plasma pTau181 (pg/ml), and plasma pTau231 (pg/ml) in the CN Aβ+ group.

Plasma sTREM2 association with brain Aβ load, AD-related biomarkers, neuroinflammation, and peripheral inflammation markers were investigated using the Spearman ation, and *p*-values < 0.05 (bold) were considered significant. Generalised linear models were utilised to explore plasma sTREM2 association with brain Aβ load, AD-d biomarkers, neuroinflammation, and peripheral inflammation markers upon adjusting for age, gender, and *APOE* ε4 status, followed by FDR correction. *n*’ represents mber of participants, ‘*ρ*’ represents the Spearman correlation coefficient and *β*’ represents the beta coefficient. Parameters significantly correlating (*p*-values < 0.05, bold) lasma sTREM2 after adjusting for confounding variables and survived FDR adjustment (*p*-values < 0.05, bold and ‘≠’ signed) were considered significant.

### Association between plasma sTREM2 with neuroinflammation and peripheral inflammation blood markers based on brain Aβ status

Neuroinflammation markers (measured with KP metabolites) and peripheral inflammation marker (measured with CRP) associated with plasma sTREM2 were investigated in all participants and separately in CN Aβ− and CN Aβ+ utilising the Spearman correlation coefficient and then generalised linear model accounting for age, sex, and *APOE* ε4 status with FDR adjustment (**Table 2**). In all participants, a significant positive correlation of plasma sTREM2 was observed with serum microglial KP metabolites (Kynurenine, 3-Hydroxykynurenine, and Quinolinic acid) both before and after adjusting for covariates (**Table 2**, **Figure 2**). While investigating separately in CN Aβ− and CN Aβ+ individuals, a significant positive correlation of plasma sTREM2 was observed with serum Kynurenine and Quinolinic acid in both groups, before and after adjusting for covariates (**Table 2**, **Figure 2**).

**Figure 2.**
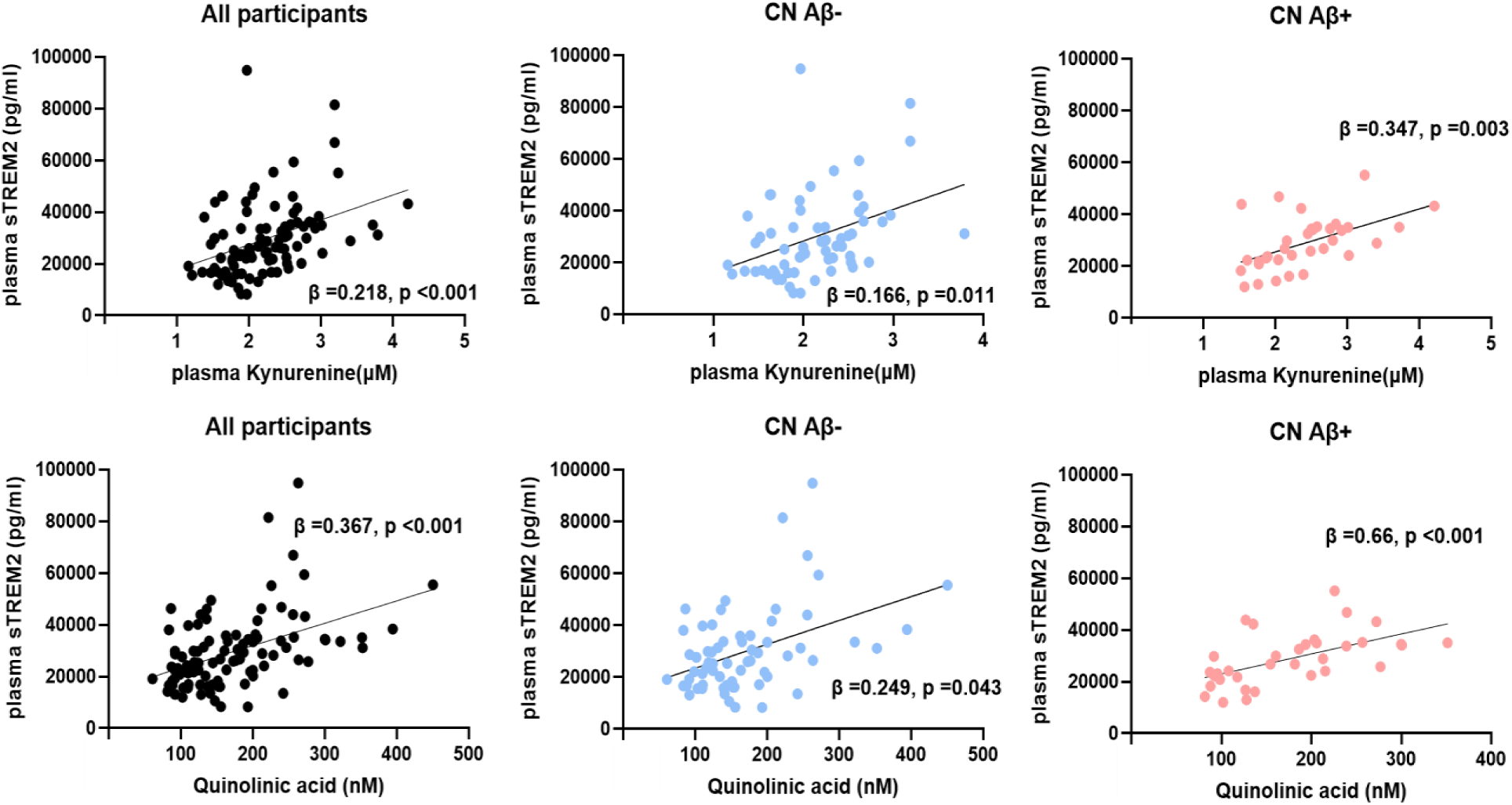
Association between plasma sTREM2 and serum microglial kynurenine pathway metabolites, irrespective of brain PET-Aβ status. (Neuro)inflammation blood biomarkers significantly correlating with plasma sTREM2 both before and after adjusting for confounding variables and survived false discovery rate correction have been reported. Plasma sTREM2 concentrations (pg/ml) positively correlated with serum Kynurenine (µM) and Quinolinic Acid (nM) in all participants and both CN Aβ- and CN Aβ+ groups.

With respect to correlation with the other serum KP metabolites that are mostly produced in the astrocytes (tryptophan, kynurenic acid, 3-Hydroxyanthranilic acid, Anthranilic acid, and Picolinic acid), there were no significant correlations observed between plasma sTREM2 after adjusting for covariates in all participants or in either CN Aβ- and CN Aβ+ groups (**Supplementary Table 1**). Additionally, there was no correlation between plasma sTREM2 and C-reactive protein (CRP) levels as peripheral markers of inflammation in general (**Table 2**).

### Influence of R47H and R62H TREM2 variants on plasma sTREM2 levels and brain A**β** Load

After stratifying the cohort based on the presence of R47H *TREM2* variant, we found heterozygous R47H *TREM2* carriers (n=5) and non-carriers (n=95) within the whole cohort. All of the five participants with heterozygous R47H carriers were of CN Aβ+ group, subjective memory complainers (SMC), and had high levels of plasma sTREM2 (**Table 3**). Of these five, three participants were *APOE* ε4 carriers and they had a higher brain Aβ load SUVR value than the two non-*APOE* ε4 carriers (participants 1-3 compared to participants 4-5, **Table 3).** After stratifying the cohort based on the presence of R62H *TREM2* variant, we found heterozygous R62H *TREM2* carriers (n=3) and non-carriers (n=97). Of these three (**Table 3)**, participant 1, an *APOE* ε4 carrier who has both R47H and R62H *TREM2* variants, showed higher brain Aβ load and plasma sTREM2 levels compared to participants 6 and 7, an *APOE* ε4 carrier with only R62H variant and non-*APOE* ε4 carrier with only R62H variant, respectively. However, brain Aβ SUVR values in R62H carriers (participants 6 and 7) were in the lower range (CN Aβ-group), suggesting the effect of *TREM2* AD rare variants on brain Aβ positivity may be attributed to R47H. To test this, we carried out the analysis solely for R47H. However, due to very low number of R62H participants, no statistical test can be performed, in particular with respect to correlation analysis ^35^.

**Table 3.** Associated factors with each R47H and R62H heterozygous carrier. ε3/ε4= APOE4 carriers; ε3/ε3=non-APOE4 carriers; Het=heterozygous TREM2 R47H/R62H carrier; NC=non-carrier of TREM2 R47H/R62H; SMC=subjective memory complainers; Y/N= yes or no, SUVR=standard uptake value ratio. Plasma sTREM2 quartiles cut-offs were 19,430.822 pg/ml, 26,777.891 pg/ml, and 35,136.831 pg/ml for Q1, Q2, and Q3, respectively.

| Participants | *APOE* | *TREM2* R47H | *TREM2* R62H | Age range (quartiles) | Gender | SMC | Brain Aβ (SUVR) | plasma sTREM2 in pg/ml (quartiles) |
| --- | --- | --- | --- | --- | --- | --- | --- | --- |
| 1 | $\epsilon 3/\epsilon 4$ | Het | Het | 79-83 (Q3) | M | Y | 1.87 | 44588.02 (>Q3) |
| 2 | $\epsilon 3/\epsilon 4$ | Het | NC | 66-74 (Q1) | F | Y | 1.97 | 46760.07 (>Q3) |
| 3 | $\epsilon 3/\epsilon 4$ | Het | NC | 79-83 (Q3) | F | Y | 1.99 | 42265.54 (>Q3) |
| 4 | $\epsilon 3/\epsilon 3$ | Het | NC | 84-89 (>Q3) | F | Y | 1.53 | 34412.41 (Q3) |
| 5 | $\epsilon 3/\epsilon 3$ | Het | NC | 74-79 (Q2) | F | Y | 1.47 | 28878.74 (Q3) |
| 6 | $\epsilon 3/\epsilon 4$ | NC | Het | 66-74 (Q1) | F | Y | 1.19 | 25309.84 (Q2) |
| 7 | $\epsilon 3/\epsilon 3$ | NC | Het | 66-74 (Q1) | F | N | 1.09 | 31370.31 (Q3) |

With respect to plasma sTREM2 levels, R47H carriers (n=5) had higher plasma sTREM2 concentrations compared to the age-, sex-, *APOE*-, and brain Aβ-matched non-R47H carriers (n=5), with (*p*=0.025, **Figure 3**) and without (*p*=0.006) adjusting for age, sex, and *APOE* ε4. A similar result was observed when comparing R47H carriers (n=5) with all of the non-R47H carriers (n=95) (with *p*=0.042, **Figure 3** and without adjusting for covariates *p*=0.020). Additionally, these five R47H carriers were at the highest risk of AD (CN Aβ+ with SMC).

**Figure 3.**
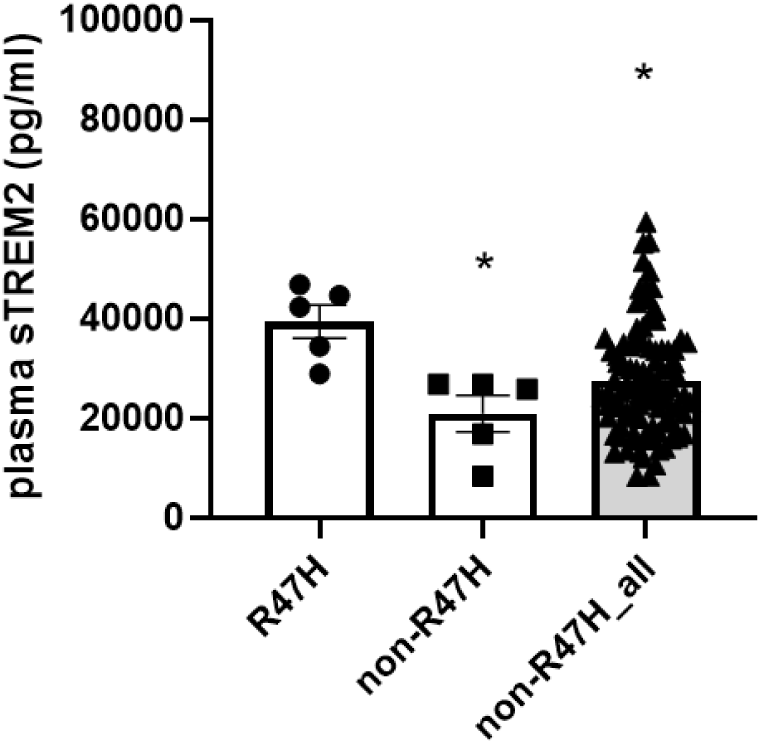
Comparison of plasma sTREM2 concentrations between R47H heterozygous carriers versus non-R47H carriers among their age-, sex-, *APOE*-, and brain PET Aβ-matched individuals, as well as among all participants. Significant increase of plasma sTREM2 concentrations (pg/ml) was observed in R47H heterozygous carriers compared to non-R47H carriers within overall participants. *p<0.05

## DISCUSSION

While CSF sTREM2 levels have been extensively studied, few studies have investigated sTREM2 in blood, and they have observed conflicting results. We aimed to investigate the levels of sTREM2 in plasma and its association with brain Aβ load and AD blood biomarkers from a cohort of well-characterized cognitively normal older adults at risk of AD, characterised with high neocortical Aβ (brain Aβ+) compared to those with low neocortical Aβ with no apparent risk of AD (brain Aβ-). Plasma sTREM2 concentrations were not significantly different in cognitively normal older adults with brain Aβ+ versus brain Aβ-participants. In line with our findings, other studies have reported no significant difference in the plasma levels of sTREM2 between AD and healthy controls ^10, 11, 21^, despite increased levels of TREM2 mRNA and blood protein observed in AD ^36^. Using an association approach, our study is the first to report a positive association between plasma sTREM2 and plasma p-tau in cognitively normal adults with high brain Aβ load (at high risk of AD). However, plasma sTREM2 levels were not correlated with brain Aβ load, indicating a positive association of plasma sTREM2 with tau-related neurodegeneration, but not with Aβ pathology in the preclinical AD stage. The present study also observed that plasma sTREM2 levels were significantly associated with neuroinflammatory marker microglial KP metabolites, such as Kynurenine and Quinolinic Acid ^37, 38^, but not KP metabolites produced mostly in the astrocytes ^39^ and classic astrocyte activation marker GFAP ^40^, indicating plasma sTREM2 levels as a marker for microglia activity in the early stage of the disease. While astrocytes primarily produce the neuroprotective Kynurenic Acid, they can also indirectly contribute to the production of the neurotoxic Quinolinic Acid, especially in the context of neuroinflammation (in the presence of activated microglia) ^41^. In accordance with the evidence which reported an inconsistent association between sTREM2 and markers of peripheral inflammation ^19^, our study showed no correlation between sTREM2 and CRP levels in the blood. In addition, R47H carriers have also shown increased levels of sTREM2 compared to non-*TREM2* carriers. Interestingly, in *APOE* ε4 carriers, the presence of R47H allele led to significantly higher levels of sTREM2 and the most pronounced brain Aβ deposition compared to R47H non-carriers (while in non-*APOE* ε4 carrier *TREM2* genotype doesn’t show any effect on sTREM2 levels).

Neuroinflammation and tau pathology are closely interconnected, which leads to AD pathobiology. Mounting evidence suggests a vicious cycle where these elements exacerbate each other ^42, 43^. Neuroinflammation, characterized by the activation of brain immune cells like microglia and astrocytes, can trigger the production and accumulation of Aβ and tau protein tangles, which are hallmarks of AD. In turn, these protein aggregates can further fuel neuroinflammation, creating a self-perpetuating cycle of damage. While CSF sTREM2 levels might serve as a biomarker for inflammatory diseases including AD, particularly as a marker for microglia activity in the early stage of the disease ^13, 14^, it is unclear whether there is a relationship between blood sTREM2 and (neuro)inflammation markers. Only one study, in people with Down Syndrome predementia has reported significantly elevated plasma sTREM2 and inflammatory markers ^44^. Interestingly, recent study has shown that plasma sTREM2-related inflammatory activity is altered in the early stages of AD ^45^. With regards to the relationship of sTREM2 to phosphorylated tau (p-tau), only CSF sTREM2 has been shown to have a close association with CSF p-tau, but not with Aβ ^17, 46^, as well as a close association between plasma sTREM2 and CSF p-tau (S199) only ^47^, suggesting the important role of sTREM2 in the development of AD pathology and tau-induced neurodegeneration.

Whether plasma sTREM2 reflects CSF sTREM2 in AD progression remains unclear, however, increases in plasma sTREM2 are reflected by increased microglial KP activity as an indicative of neuroinflammation ^31^, albeit independent of brain Aβ status. Thus, elevated sTREM2, either in the CNS or the periphery, likely represents microglial activation, but whether such innate immune activation is beneficial or detrimental throughout the course of AD progression remains unclear.

In accordance with an absence of ApoE-specific isoform-related effect on sTREM2 ^49,50^, in our study sTREM2 levels were not significantly different in *APOE* ε4 carriers vs non-carriers. However, the presence of a rare variant in the in the *TREM2* gene, R47H, has been shown to strongly affect sTREM2 levels in CSF ^10^, but did not affect sTREM2 expression in the brain of AD individuals ^48^, while the same results in serum/plasma are more elusive ^11^. This R47H variant has been associated with AD risk to a degree similar to the presence of the *APOE* ε4 allele, although it gained far less attention due to its extremely low frequency in the general population ^49, 50^. As the disease progresses, others have reported that AD individuals harboring the R47H variant of *TREM2* display no change in CSF Aβ42 levels but had increased levels of both total tau and phosphorylated tau (Thr181) compared to non-carriers ^51, 52^. However, this positive association between R47H with brain Aβ load as measured by PiB-PET imaging was no longer observed at a later stage of AD progression ^33^, suggesting the role of TREM2 in AD trajectory as a function of its interaction with tau. Thus, the exact mechanism(s) by which TREM2 affects late-onset AD risk remains to be determined.

It has been reported that R62H *TREM2* variant may also play a role in AD risk ^53, 54^, albeit with a lower impact on lipid binding than the R47H variant ^55^. Interestingly, sTREM2 derived from these AD-associated variants, R47H and R62H, are less potent in both suppressing apoptosis and triggering inflammatory responses in microglia ^56^. However, unlike R47H, the R62H *TREM2* variant is associated with reduced or unchanged CSF and brain sTREM2 levels in AD individuals ^10, 20^, albeit it increased the full-length TREM2 expression in the brain ^5^. Even though the number of people with R62H is too small in our study, there is an indication that people harboring R62H variant had lower plasma sTREM2 levels and brain Aβ load compared to R47H carriers.

The current study on investigating plasma sTREM2 for AD-related clinical significance has many strengths. Firstly, it utilizes a highly characterized, cognitively normal cohort with a representative proportion of preclinical AD individuals, in agreement with other established cohorts ^57, 58^, employing PET for brain Aβ SUVR measurement, a stronger marker of AD neuropathology compared to CSF Aβ. Secondly, the study incorporates a highly sensitive assay to measure plasma sTREM2. We acknowledge, however, that the current study has limitations regarding its relatively modest sample size and cross-sectional design. In addition, given the low frequency of R47H *TREM2* variant in our study and other studies ^49,50,59^ and the small sample size for R47H and R62H *TREM2* genetic variant carriers, these variants might possess minimal clinical utility as a predictor or diagnostic for AD. Therefore, further studies are required to validate the current findings in larger independent cohorts, using both cross-sectional and longitudinal study designs. Longitudinal studies will provide more insight into the trajectory of plasma sTREM2 alterations associated with the progression of AD pathogenesis.

## CONCLUSION

The major findings of the study are that positive correlations exist between plasma sTREM2 concentrations with plasma p-tau (181 and 231) concentrations in older adults at high risk of AD. Our data indicated that plasma sTREM2 may serve as a potentially useful biomarker for microglia activity in the early stage of AD and consequent neuroinflammation in the AD continuum. Furthermore, R47H rare variants of *TREM2* may affect brain Aβ deposition at the early stage of AD, as indicated by increased plasma sTREM2 and increased brain Aβ positivity in cognitively normal adults at high risk of AD harboring R47H *TREM2* alleles.

## Data Availability

All data produced in the present study are available upon reasonable request to the authors

## Author Contributions

Prita R. Asih (Conceptualization; Formal analysis; Investigation, Resources; Data Curation; Writing-Original Draft; Writing-Review & Editing, Visualization), Cameron W. Morris (Methodology; Software; Validation; Writing-Review & Editing), Hong Wang (Methodology; Software; Validation; Supervision), Steve Pedrini (Formal analysis; Writing-Review & Editing), Kathryn Goozee (Resources; Investigation; Project administration; Writing-Review & Editing), Kevin Taddei (Resources; Investigation; Project administration; Writing-Review & Editing), Hamid R. Sohrabi (Investigation; Writing-Review & Editing), Simon M. Law (Resources; Investigation; Writing-Review & Editing), Pratishtha Chatterjee (Resources; Investigation; Writing-Review & Editing), Stephanie R. Rainey-Smith (Investigation; Writing-Review & Editing), Chai K Lim (Methodology; Writing-Review & Editing), Gilles J. Guillemin (Resources; Methodology; Writing-Review & Editing), Thomas J. Karikari (Resources; Methodology; Writing-Review & Editing), Colin L. Masters (Investigation; Writing-Review & Editing), Henrik Zetterberg (Investigation; Writing-Review & Editing), Kaj Blennow (Investigation; Writing-Review & Editing), Nicholas J. Ashton (Investigation, Writing-Review & Editing), Ralph N. Martins (Conceptualization; Writing-Review & Editing; Supervision; Project administration; Funding acquisition).

## Acknowledgments

We thank the participants and their families for their participation and cooperation, and the Anglicare, KaRa Minds and Alzheimer’s Research Australia (ARA) research and support staff for their contributions to this study. We also thank the staff of the Macquarie Medical Imaging centre in Macquarie University Hospital, Sydney, for their contributions. Florbetaben is a proprietary PET radiopharmaceutical owned by Piramal Imaging. For this study, Florbetaben was manufactured and supplied under GMP conditions by Cyclotek (Australia) Pty Ltd.

## Funding

The authors have been supported by grants from Lions Alzheimer’s Research Foundation (LAF) and Alzheimer’s Research Australia (ARA).

## Declaration of conflicting interests

The authors declared no potential conflicts of interest with respect to the research, authorship, and/or publication of this article.

***Datasets/Data Availability Statement (Required for Research Reports, Short Communications, and Systematic Reviews/Meta-Analyses)***

The data supporting the findings of this study are available on request from the corresponding author. The data are not publicly available due to privacy or ethical restrictions.

## Notes

### Competing Interest Statement

The authors have declared no competing interest.

### Funding Statement

The study have been supported by grants from Lions Alzheimers Research Foundation (LAF) and Alzheimers Research Australia (ARA).

### Author Declarations

The Bellberry Human Research Ethics Committee, Australia, provided ethical approval for the study

## REFERENCES

1. Jonsson T, Stefansson H, Steinberg S, et al. Variant of TREM2 associated with the risk of Alzheimer’s disease. N Engl J Med 2013; 368: 107–116. 20121114. DOI: 10.1056/NEJMoa1211103.

2. Guerreiro R, Wojtas A, Bras J, et al. TREM2 variants in Alzheimer’s disease. N Engl J Med 2013; 368: 117–127. 20121114. DOI: 10.1056/NEJMoa1211851.

3. Carmona S, Zahs K, Wu E, et al. The role of TREM2 in Alzheimer’s disease and other neurodegenerative disorders. Lancet Neurol 2018; 17: 721–730. 20180717. DOI: 10.1016/S1474-4422(18)30232-1.

4. Song WM, Joshita S, Zhou Y, et al. Humanized TREM2 mice reveal microglia-intrinsic and - extrinsic effects of R47H polymorphism. J Exp Med 2018; 215: 745–760. 20180110. DOI: 10.1084/jem.20171529.

5. Sims R, van der Lee SJ, Naj AC, et al. Rare coding variants in PLCG2, ABI3, and TREM2 implicate microglial-mediated innate immunity in Alzheimer’s disease. Nat Genet 2017; 49: 1373–1384. 20170717. DOI: 10.1038/ng.3916.

6. Korvatska O, Leverenz JB, Jayadev S, et al. R47H Variant of TREM2 Associated With Alzheimer Disease in a Large Late-Onset Family: Clinical, Genetic, and Neuropathological Study. JAMA Neurol 2015; 72: 920–927. DOI: 10.1001/jamaneurol.2015.0979.

7. Yeh FL, Hansen DV and Sheng M. TREM2, Microglia, and Neurodegenerative Diseases. Trends Mol Med 2017; 23: 512–533. 2017/04/27. DOI: 10.1016/j.molmed.2017.03.008.

8. Boza-Serrano A, Ruiz R, Sanchez-Varo R, et al. Galectin-3, a novel endogenous TREM2 ligand, detrimentally regulates inflammatory response in Alzheimer’s disease. Acta Neuropathol 2019; 138: 251–273. 2019/04/22. DOI: 10.1007/s00401-019-02013-z.

9. Zhong L, Wang Z, Wang D, et al. Amyloid-beta modulates microglial responses by binding to the triggering receptor expressed on myeloid cells 2 (TREM2). Mol Neurodegener 2018; 13: 15. 2018/03/29. DOI: 10.1186/s13024-018-0247-7.

10. Piccio L, Deming Y, Del-Aguila JL, et al. Cerebrospinal fluid soluble TREM2 is higher in Alzheimer disease and associated with mutation status. Acta Neuropathol 2016; 131: 925–933. 20160111. DOI: 10.1007/s00401-016-1533-5.

11. Ashton NJ, Suarez-Calvet M, Heslegrave A, et al. Plasma levels of soluble TREM2 and neurofilament light chain in TREM2 rare variant carriers. Alzheimers Res Ther 2019; 11: 94. 20191128. DOI: 10.1186/s13195-019-0545-5.

12. Wunderlich P, Glebov K, Kemmerling N, et al. Sequential proteolytic processing of the triggering receptor expressed on myeloid cells-2 (TREM2) protein by ectodomain shedding and gamma-secretase-dependent intramembranous cleavage. J Biol Chem 2013; 288: 33027–33036. 20130927. DOI: 10.1074/jbc.M113.517540.

13. Suarez-Calvet M, Kleinberger G, Araque Caballero MA, et al. sTREM2 cerebrospinal fluid levels are a potential biomarker for microglia activity in early-stage Alzheimer’s disease and associate with neuronal injury markers. EMBO Mol Med 2016; 8: 466–476. 2016/03/05. DOI: 10.15252/emmm.201506123.

14. Rauchmann BS, Sadlon A, Perneczky R, et al. Soluble TREM2 and Inflammatory Proteins in Alzheimer’s Disease Cerebrospinal Fluid. J Alzheimers Dis 2020; 73: 1615–1626. 2020/01/21. DOI: 10.3233/JAD-191120.

15. Wang R, Zhan Y, Zhu W, et al. Association of soluble TREM2 with Alzheimer’s disease and mild cognitive impairment: a systematic review and meta-analysis. Front Aging Neurosci 2024; 16: 1407980. 2024/06/06. DOI: 10.3389/fnagi.2024.1407980.

16. Henjum K, Almdahl IS, Arskog V, et al. Cerebrospinal fluid soluble TREM2 in aging and Alzheimer’s disease. Alzheimers Res Ther 2016; 8: 17. 2016/04/29. DOI: 10.1186/s13195-016-0182-1.

17. Suarez-Calvet M, Araque Caballero MA, Kleinberger G, et al. Early changes in CSF sTREM2 in dominantly inherited Alzheimer’s disease occur after amyloid deposition and neuronal injury. Sci Transl Med 2016; 8: 369ra178. 2016/12/16. DOI: 10.1126/scitranslmed.aag1767.

18. Morenas-Rodriguez E, Li Y, Nuscher B, et al. Soluble TREM2 in CSF and its association with other biomarkers and cognition in autosomal-dominant Alzheimer’s disease: a longitudinal observational study. Lancet Neurol 2022; 21: 329–341. 2022/03/20. DOI: 10.1016/S1474-4422(22)00027-8.

19. Heslegrave A, Heywood W, Paterson R, et al. Increased cerebrospinal fluid soluble TREM2 concentration in Alzheimer’s disease. Mol Neurodegener 2016; 11: 3. 2016/01/13. DOI: 10.1186/s13024-016-0071-x.

20. Suarez-Calvet M, Morenas-Rodriguez E, Kleinberger G, et al. Early increase of CSF sTREM2 in Alzheimer’s disease is associated with tau related-neurodegeneration but not with amyloid-beta pathology. Mol Neurodegener 2019; 14: 1. 2019/01/12. DOI: 10.1186/s13024-018-0301-5.

21. Du SH, Zhang W, Yue X, et al. Role of CXCR1 and Interleukin-8 in Methamphetamine-Induced Neuronal Apoptosis. Front Cell Neurosci 2018; 12: 230. 2018/08/21. DOI: 10.3389/fncel.2018.00230.

22. Ohara T, Hata J, Tanaka M, et al. Serum Soluble Triggering Receptor Expressed on Myeloid Cells 2 as a Biomarker for Incident Dementia: The Hisayama Study. Ann Neurol 2019; 85: 47–58. 2018/11/30. DOI: 10.1002/ana.25385.

23. Schulz I, Kruse N, Gera RG, et al. Systematic Assessment of 10 Biomarker Candidates Focusing on alpha-Synuclein-Related Disorders. Mov Disord 2021; 36: 2874–2887. 2021/08/08. DOI: 10.1002/mds.28738.

24. Goozee K, Chatterjee P, James I, et al. Alterations in erythrocyte fatty acid composition in preclinical Alzheimer’s disease. Sci Rep 2017; 7: 676. 2017/04/08. DOI: 10.1038/s41598-017-00751-2.

25. Nasreddine ZS, Phillips N and Chertkow H. Normative data for the Montreal Cognitive Assessment (MoCA) in a population-based sample. Neurology 2012; 78: 765–766; author reply 766. 2012/03/07. DOI: 10.1212/01.wnl.0000413072.54070.a3.

26. McKhann GM, Knopman DS, Chertkow H, et al. The diagnosis of dementia due to Alzheimer’s disease: recommendations from the National Institute on Aging-Alzheimer’s Association workgroups on diagnostic guidelines for Alzheimer’s disease. Alzheimers Dement 2011; 7: 263–269. 2011/04/26. DOI: 10.1016/j.jalz.2011.03.005.

27. Folstein MF, Folstein SE and McHugh PR. “Mini-mental state”. A practical method for grading the cognitive state of patients for the clinician. J Psychiatr Res 1975; 12: 189–198. 1975/11/01. DOI: 10.1016/0022-3956(75)90026-6.

28. Zhou L, Salvado O, Dore V, et al. MR-less surface-based amyloid assessment based on 11C PiB PET. PLoS One 2014; 9: e84777. 2014/01/16. DOI: 10.1371/journal.pone.0084777.

29. Bourgeat P, Villemagne VL, Dore V, et al. Comparison of MR-less PiB SUVR quantification methods. Neurobiol Aging 2015; 36 Suppl 1: S159–166. 2014/09/27. DOI: 10.1016/j.neurobiolaging.2014.04.033.

30. Goozee K, Chatterjee P, James I, et al. Elevated plasma ferritin in elderly individuals with high neocortical amyloid-beta load. Mol Psychiatry 2018; 23: 1807–1812. 20170711. DOI: 10.1038/mp.2017.146.

31. Chatterjee P, Goozee K, Lim CK, et al. Alterations in serum kynurenine pathway metabolites in individuals with high neocortical amyloid-beta load: A pilot study. Sci Rep 2018; 8: 8008. 20180522. DOI: 10.1038/s41598-018-25968-7.

32. Chatterjee P, Goozee K, Sohrabi HR, et al. Association of Plasma Neurofilament Light Chain with Neocortical Amyloid-beta Load and Cognitive Performance in Cognitively Normal Elderly Participants. J Alzheimers Dis 2018; 63: 479–487. DOI: 10.3233/JAD-180025.

33. Rosenthal SL, Bamne MN, Wang X, et al. More evidence for association of a rare TREM2 mutation (R47H) with Alzheimer’s disease risk. Neurobiol Aging 2015; 36: 2443 e2421–2446. 20150425. DOI: 10.1016/j.neurobiolaging.2015.04.012.

34. Chatterjee P, Pedrini S, Ashton NJ, et al. Diagnostic and prognostic plasma biomarkers for preclinical Alzheimer’s disease. Alzheimers Dement 2022; 18: 1141–1154. 20210908. DOI: 10.1002/alz.12447.

35. Aggarwal R and Ranganathan P. Common pitfalls in statistical analysis: The use of correlation techniques. Perspect Clin Res 2016; 7: 187–190. 2016/11/16. DOI: 10.4103/2229-3485.192046.

36. Hu N, Tan MS, Yu JT, et al. Increased expression of TREM2 in peripheral blood of Alzheimer’s disease patients. J Alzheimers Dis 2014; 38: 497–501. 2013/09/05. DOI: 10.3233/JAD-130854.

37. Guillemin GJ. Quinolinic acid, the inescapable neurotoxin. FEBS J 2012; 279: 1356–1365. 20120327. DOI: 10.1111/j.1742-4658.2012.08485.x.

38. Garrison AM, Parrott JM, Tunon A, et al. Kynurenine pathway metabolic balance influences microglia activity: Targeting kynurenine monooxygenase to dampen neuroinflammation. Psychoneuroendocrinology 2018; 94: 1–10. 20180422. DOI: 10.1016/j.psyneuen.2018.04.019.

39. Cortes Malagon EM, Lopez Ornelas A, Olvera Gomez I, et al. The Kynurenine Pathway, Aryl Hydrocarbon Receptor, and Alzheimer’s Disease. Brain Sci 2024; 14 20240923. DOI: 10.3390/brainsci14090950.

40. Liddelow SA and Barres BA. Reactive Astrocytes: Production, Function, and Therapeutic Potential. Immunity 2017; 46: 957-967. DOI: 10.1016/j.immuni.2017.06.006.

41. Ting KK, Brew B and Guillemin G. The involvement of astrocytes and kynurenine pathway in Alzheimer’s disease. Neurotox Res 2007; 12: 247–262. DOI: 10.1007/BF03033908.

42. Kiraly M, Foss JF and Giordano T. Neuroinflammation, its Role in Alzheimer’s Disease and Therapeutic Strategie. J Prev Alzheimers Dis 2023; 10: 686–698. DOI: 10.14283/jpad.2023.109.

43. Novoa C, Salazar P, Cisternas P, et al. Inflammation context in Alzheimer’s disease, a relationship intricate to define. Biol Res 2022; 55: 39. 20221223. DOI: 10.1186/s40659-022-00404-3.

44. Weber GE, Koenig KA, Khrestian M, et al. An Altered Relationship between Soluble TREM2 and Inflammatory Markers in Young Adults with Down Syndrome: A Preliminary Report. J Immunol 2020; 204: 1111–1118. 2020/01/22. DOI: 10.4049/jimmunol.1901166.

45. Weber GE, Khrestian M, Tuason ED, et al. Peripheral sTREM2-Related Inflammatory Activity Alterations in Early-Stage Alzheimer’s Disease. J Immunol 2022; 208: 2283–2299. 2022/05/07. DOI: 10.4049/jimmunol.2100771.

46. Deczkowska A, Weiner A and Amit I. The Physiology, Pathology, and Potential Therapeutic Applications of the TREM2 Signaling Pathway. Cell 2020; 181: 1207–1217. 2020/06/13. DOI: 10.1016/j.cell.2020.05.003.

47. Park SH, Lee EH, Kim HJ, et al. The relationship of soluble TREM2 to other biomarkers of sporadic Alzheimer’s disease. Sci Rep 2021; 11: 13050. 2021/06/24. DOI: 10.1038/s41598-021-92101-6.

48. Ma L, Allen M, Sakae N, et al. Expression and processing analyses of wild type and p.R47H TREM2 variant in Alzheimer’s disease brains. Mol Neurodegener 2016; 11: 72. 2016/11/27. DOI: 10.1186/s13024-016-0137-9.

49. Hooli BV, Parrado AR, Mullin K, et al. The rare TREM2 R47H variant exerts only a modest effect on Alzheimer disease risk. Neurology 2014; 83: 1353–1358. 2014/09/05. DOI: 10.1212/WNL.0000000000000855.

50. Zhou SL, Tan CC, Hou XH, et al. TREM2 Variants and Neurodegenerative Diseases: A Systematic Review and Meta-Analysis. J Alzheimers Dis 2019; 68: 1171–1184. 2019/03/19. DOI: 10.3233/JAD-181038.

51. Lill CM, Rengmark A, Pihlstrom L, et al. The role of TREM2 R47H as a risk factor for Alzheimer’s disease, frontotemporal lobar degeneration, amyotrophic lateral sclerosis, and Parkinson’s disease. Alzheimers Dement 2015; 11: 1407–1416. 2015/05/06. DOI: 10.1016/j.jalz.2014.12.009.

52. Cruchaga C, Kauwe JS, Harari O, et al. GWAS of cerebrospinal fluid tau levels identifies risk variants for Alzheimer’s disease. Neuron 2013; 78: 256–268. 2013/04/09. DOI: 10.1016/j.neuron.2013.02.026.

53. Jin SC, Benitez BA, Karch CM, et al. Coding variants in TREM2 increase risk for Alzheimer’s disease. Hum Mol Genet 2014; 23: 5838–5846. 2014/06/06. DOI: 10.1093/hmg/ddu277.

54. Cuyvers E, Bettens K, Philtjens S, et al. Investigating the role of rare heterozygous TREM2 variants in Alzheimer’s disease and frontotemporal dementia. Neurobiol Aging 2014; 35: 726 e711–729. 2013/10/15. DOI: 10.1016/j.neurobiolaging.2013.09.009.

55. Yeh FL, Wang Y, Tom I, et al. TREM2 Binds to Apolipoproteins, Including APOE and CLU/APOJ, and Thereby Facilitates Uptake of Amyloid-Beta by Microglia. Neuron 2016; 91: 328–340. 2016/08/02. DOI: 10.1016/j.neuron.2016.06.015.

56. Zhong L, Chen XF, Wang T, et al. Soluble TREM2 induces inflammatory responses and enhances microglial survival. J Exp Med 2017; 214: 597–607. 2017/02/18. DOI: 10.1084/jem.20160844.

57. Ellis KA, Bush AI, Darby D, et al. The Australian Imaging, Biomarkers and Lifestyle (AIBL) study of aging: methodology and baseline characteristics of 1112 individuals recruited for a longitudinal study of Alzheimer’s disease. Int Psychogeriatr 2009; 21: 672–687. 2009/05/28. DOI: 10.1017/S1041610209009405.

58. Mueller SG, Weiner MW, Thal LJ, et al. The Alzheimer’s disease neuroimaging initiative. Neuroimaging Clin N Am 2005; 15: 869–877, xi-xii. 2006/01/31. DOI: 10.1016/j.nic.2005.09.008.

59. Ayer AH, Wojta K, Ramos EM, et al. Frequency of the TREM2 R47H Variant in Various Neurodegenerative Disorders. Alzheimer Dis Assoc Disord 2019; 33: 327–330. DOI: 10.1097/WAD.0000000000000339.

