## Supplemental Figure and Table for "Plasma soluble TREM2 is associated with plasma pTau-181 and pTau-231 in cognitively normal older adults at risk of Alzheimer’s disease"

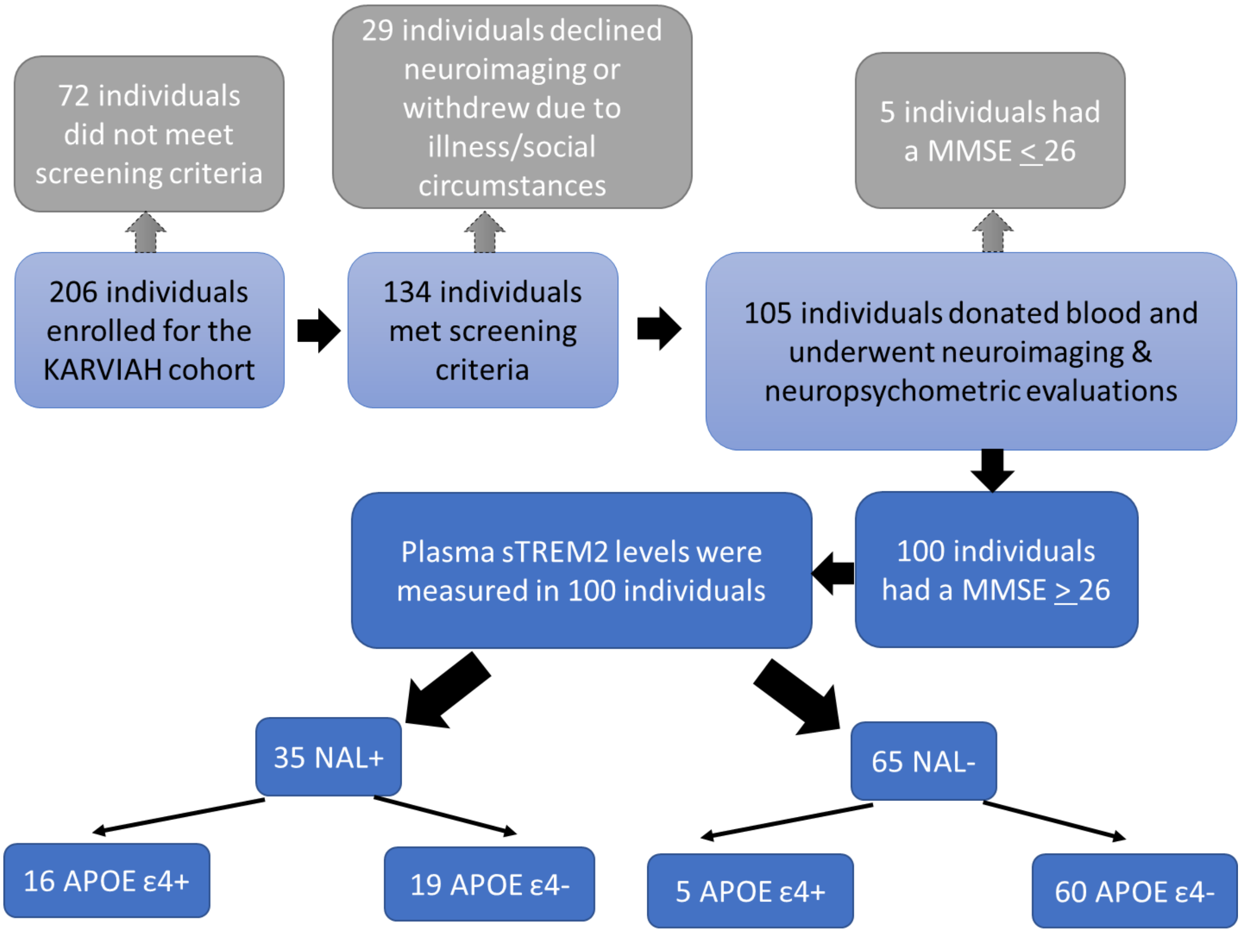


**Supplement Figure 1. Flowchart illustrating participants included within the current study.**

**Table S1. Plasma sTREM2 association with serum non-microglia specific kynurenine pathway (KP) metabolites in all participants, CN Aβ- and CN Aβ+**

| | All participants (n=100) | | | | | | CN Aβ- (n=65) | | | | | | CN Aβ+ (n=35) | | | | | |
|---|---|---|---|---|---|---|---|---|---|---|---|---|---|---|---|---|---|---|
| | sTREM2 | | | | | | sTREM2 | | | | | | sTREM2 | | | | | |
| | *p* | p-value | 95% CI | β | p-value | 95% CI | *p* | p-value | 95% CI | β | p-value | 95% CI | *p* | p-value | 95% CI | β | p-value | 95% CI |
| Tryptophan (µM) | -0.018 | 0.86 | 221-0.187 | -0.003 | 0.932 | -0.083-0.077 | -0.095 | 0.457 | 339-0.162 | -0.01 | 0.833 | -0.102-0.082 | 0.105 | 0.555 | 243-0.427 | 0.043 | 0.89 | -0.11-0.196 |
| Kynurenic acid (nM) | 0.256 | **0.011** | 055-0.438 | 0.116 | 0.089 | 0.003-0.229 | 0.372 | **0.002** | 132-0.571 | 0.254 | 0.172 | 0.077-0.43 | 0.233 | 0.185 | 124-0.537 | 0.218 | 0.732 | -0.189-0.625 |
| 3-Hydroxyanthranilic acid (nM) | 0.007 | 0.945 | 197-0.211 | -0.008 | 0.938 | -0.187-0.171 | 0.01 | 0.934 | 243-0.263 | 0.011 | 0.914 | -0.19-0.212 | -0.053 | 0.765 | -0.393-0.3 | -0.094 | 0.792 | -0.456-0.267 |
| Anthranilic acid (nM) | 0.063 | 0.535 | 143-0.264 | 0.002 | 0.987 | -0.202-0.206 | 0.111 | 0.385 | 146-0.353 | 0.077 | 0.621 | -0.13-0.284 | -0.008 | 0.964 | ).354-0.34 | -0.299 | 0.264 | -0.739-0.141 |
| Picolinic acid (nM) | -0.037 | 0.72 | 239-0.169 | -0.047 | 0.745 | -0.201-0.107 | -0.125 | 0.325 | 366-0.132 | -0.138 | 0.401 | -0.323-0.047 | 0.13 | 0.464 | 228-0.457 | 0.184 | 0.607 | -0.088-0.456 |

*Note*: Plasma sTREM2 association with serum KP metabolites as neuroinflammation markers were investigated using the Spearman correlation, and *p*-values < 0.05 (bold) were considered significant. Generalised linear models were utilised to explore plasma sTREM2 association with KP metabolites upon adjusting for age, gender, and *APOE* ε4 status, followed by FDR correction. *n*' represents the number of participants, '*ρ*' represents the Spearman correlation coefficient and *β*' represents the beta coefficient. Parameters significantly correlating (*p*-values < 0.05, bold) with plasma sTREM2 after adjusting for confounding variables and survived FDR adjustment (*p*-values < 0.05, bold and '≠' signed) were considered significant.
